# Predicting congenital syphilis: Using machine learning to enhance disease management and control

**DOI:** 10.1101/2024.04.11.24305694

**Authors:** Élisson da Silva Rocha, Cleber Matos de Morais, Igor Vitor Teixeira, Waldemar Brandão Neto, Theo Lynn, Patricia Takako Endo

## Abstract

**Objective:** Sexually Transmitted Infections (STIs) present significant challenges to global public health, affecting physical and mental well-being and straining healthcare systems and economies. This study aims to enhance the predictive performance of models for congenital syphilis prediction by incorporating additional information obtained during gestational follow-up. Building upon the work of Teixeira et al. [1], which utilizes clinical and sociodemographic data, our model was enriched with results from venereal disease research laboratory (VDRL) and rapid tests for congenital syphilis conducted on pregnant women.

**Method:** The dataset utilized in this study comprised 47,604 records spanning the period from 2013 to 2022, with 27 attributes collected from pregnant women enrolled in the Mãe Coruja Pernambucana Program in Pernambuco, Brazil. Among these attributes, we included clinical and sociodemographic factors, as well as results from venereal disease research laboratory (VDRL) and rapid tests for congenital syphilis.

**Results:** Our proposed model surpassed Teixeira’s models exhibiting higher specificity (94.74%) and a slight increase in sensitivity (70.37%).

**Conclusions:** Our study highlights the value of incorporating additional information from VDRL and rapid tests into models for predicting congenital syphilis. The combined approach involving both clinical, sociodemographic, and test result data enhances the accuracy of predictions thereby facilitating better informed healthcare decisions at different stages of pregnancy. This approach also holds significant potential in combating and managing congenital syphilis by providing assistance to health system decision makers and public policymakers. As a result, it can ultimately enhance the overall outcomes of maternal and child health and contribute to disease control.

## Introduction

The World Health Organization (WHO) [2] estimates that over 7 million people worldwide were infected by syphilis in 2020. Syphilis is a curable and treatable disease caused by the bacterium Treponema pallidum [3]. It is a complex infection with diverse clinical manifestations and three stages [3–5]. Syphilis during pregnancy is the second leading cause of stillbirths globally, and also contributes to preterm birth, low birth weight, neonatal death, and infections in newborns, amongst other conditions [4]. Congenital syphilis (CS) is the condition where syphilis is transmitted from an infected mother to her baby during pregnancy or delivery; it can lead to severe sequelae including stillbirth, neonatal death, low-birth weight and prematurity, sepsis, neonatal conjunctivitis and congenital deformities [6, 7].

With 22,065 reported cases of CS in 2020, Brazil has the highest number of reported cases of CS in the Americas [5]. Studies have consistently note that high incidences of CS in the north and northeast regions of Brazil, with the Brazilian state capitals of Recife, Campo Grande, Rio de Janeiro, Porto Alegre, and Manaus having the greatest spatial and spatiotemporal CS risk [8, 9]. Changes in migration patterns, the structure of regional health care services, delayed treatment, the prevalence of untreated partners, and penicillin shortages are amongst the factors attributed to the increased CS incidences and CS risk in these regions [8–10].

Machine learning holds great promise in transforming STIs surveillance and interventions [11]. Recently, Teixeira et al. [1] assessed the efficacy of various machine learning models in predicting adverse outcomes associated with CS using data collected from 2013 to 2021 by Mãe Coruja Pernambucana Program (PMCP), a social program in Pernambuco, Brazil. The PMCP data included clinical and sociodemographic data regarding antenatal care and the outcomes for pregnant women and their children [1]. The pre-processed data set used by Teixeira et al. [1] is publicly available at https://data.mendeley.com/datasets/3zkcvybvkz/1. Despite promising results, none of the models evaluated in their work achieved an accuracy greater than 70%, illustrating the difficulty in classifying possible outcomes of CS using only clinical and sociodemographic data [1].

Our main objective is to examine whether the inclusion of additional information acquired during the gestational follow-up process leads to an improvement in the predictive performance of models for CS. We supplement the clinical and sociodemographic data used by Teixeira et al [1] with venereal disease research laboratory (VDRL) and rapid test results from pregnant women. By incorporating this extra information for training our models, our hypothesis is that we can improve the model prediction process and, consequently, may be able to provide more precise predictions.

## Materials and methods

### Data set

We declare that the research has been approved by the Brazilian Human Research Ethics Board (Comitê de Ética em Pesquisa [CEP]) under number 12438019.2.0000.5208 and all methods were performed in accordance with the Brazilian regulations that do not require consent for studies using unidentified data from the Brazilian data health systems.

Similar to Teixeira et al. [1], the data used in this work was provided by the PMCP. The data that support the findings of this study are openly available in Mendeley Data at http://doi.org/10.17632/3zkcvybvkz.2. It comprises 12 anonymized tables extracted from the Sistema de Informação Mãe Coruja (SIS-MC) covering the period from 2013 to 2022; Teixeira et al. [1] use data up to 2021. Each table is related to a specific type of information including child-related information, childbirth, gestation, prenatal care, women’s profiles, and other relevant information related to pregnancy and child health, that were collected during prenatal care appointments.

After completing the tables merging process, our data set was consolidated into a unified data set consisting of 256 attributes and 218,014 records. In this data set, we identified 1,003 records as positive for CS and 46,666 as negative. It is important to note that over 170,000 records (78% of the data set) do not contain any information on CS. These empty records represent a major challenge for the SIS-MC, as they may indicate that tests are not being carried out right after childbirth or that the process of digitizing information is not being done appropriately. As it is not possible to describe all 256 original attributes, it is possible to find the data dictionary at: https://www.dropbox.com/scl/fi/3zbenbf6jhkqme0yoz725/Attributes-dicionary.pdf?rlkey=fddwiyr2zs8tiecfpsybdkjnp&dl=0.

### Data pre-processing

In order to use this unified data set to train our machine learning models, we perform some important pre-processing tasks including data cleaning and transformation that assist in the process of training and evaluating the models. Figure 1 illustrates the steps used to pre-process our data set.

**Fig 1.**
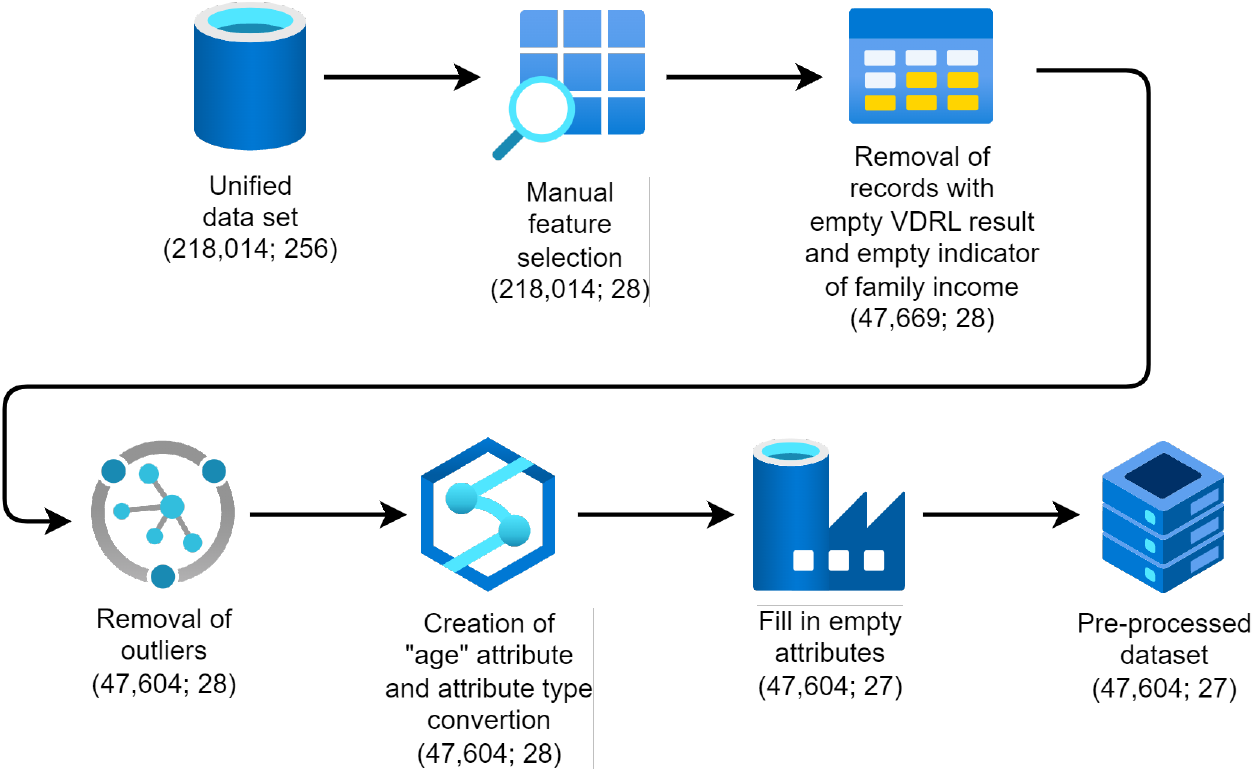
Data pre-processing methodology (Adapted from [1]).

The pre-processing methodology used in this study was adapted from Teixeira et al. [1] as we intend to compare our results with theirs. The same five steps were performed: a) manual feature selection; b) removal of records with empty indicators of family income; c) removal of outliers; d) creation of the “age” attribute and attribute type conversion; and e) filling in empty attributes. These steps are further detailed in Teixeira et al. [1]. In contrast to Teixeira et al. [1], we added a new attribute (labeled Mother_Syphilis_Test_Result), the focus of our study.

The number of records in the pre-processed data set used in our study uses an updated data set with new records from 2022. This increases the total data set from 41,762 records as per Teixeira et al. [1] to 47,604 records in this study. This increase also led to an increase in the number of positive cases for CS from 826 in Teixeira et al. [1] to 1002 in this study.

### Experiments’ methodology

Our objective is to investigate whether the inclusion of information obtained during pregnancy can improve the performance of our models in predicting CS. Our hypothesis is that the addition of the attribute related to the first syphilis test performed on the mother during pregnancy (Mother_Syphilis_Test_Result) impacts the model’s learning and, consequently, its performance in the evaluation metrics.

For comparison, we use the scenario that produced the best results by Teixeira et al. [1]. This scenario, called BODS-Expert (Balanced with One-hot Encoding Data Set from the expert), which is a data set built by considering attributes carefully chosen by health experts with balanced data, applying the one-hot encoding technique.

In this study, we want to compare and discuss the performance of our proposed approach (named **Silva scenario**) against two other approaches. The Silva scenario involves replicating the data set with the inclusion of the attribute indicating the first syphilis test performed by the mother during her pregnancy (Mother_Syphilis_Test_Result) and training a set of machine learning models. The other approaches are the **Original Teixeira model** and the **Updated Teixeira model**. The Original Teixeira model is the best model presented in Teixeira et al. [1]. The Updated Teixeira model replicates the scenario conducted by Teixeira et al. [1] using the same updated data set we are using in this study.

To enable the comparison, we performed additional pre-processing steps, as shown in Figure 2, to align with the Teixeira scenario and replicate the data set to retrain the Original Teixeira model and apply the same experiment in the Silva scenario with our updated data set.

**Fig 2.**
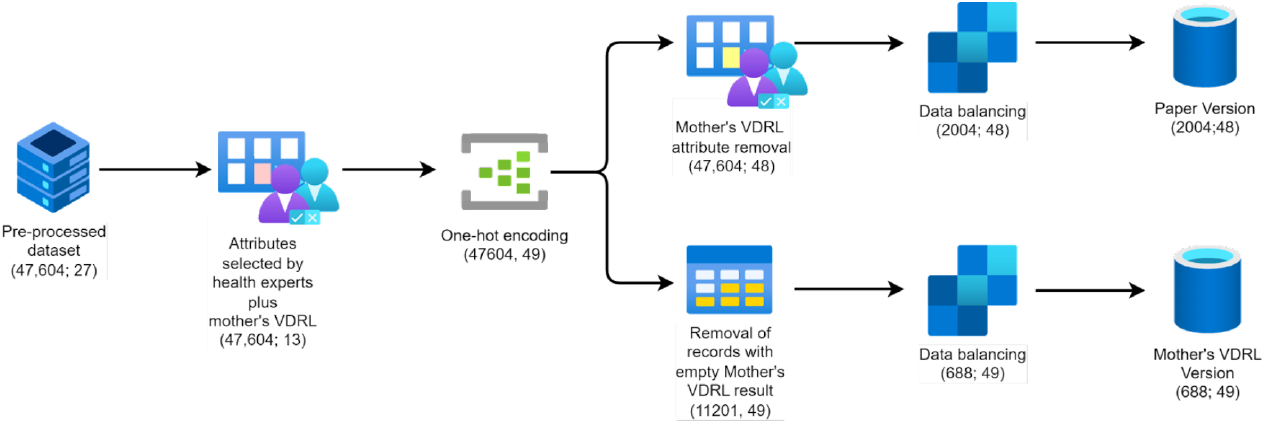
Data set replication process used for Updated Teixeira model and Silva model.

Table 1 shows the attributes selected by the health experts including the Mother_VDRL_Result attribute, comprising 13 attributes. The results of the application of one-hot encoding, transforming these 13 attributes into 49 attributes. This relates to the first two steps of Figure 2.

**Table 1.** Attributes selected by health experts plus Mother_VDRL_Result

| Attribute | Description | Type |
| --- | --- | --- |
| VDRL_Result | VDRL exam result | Binary |
| Plan_Pregnancy | Planned pregnancy | Categorical |
| Has_Preg_Risk | Has pregnancy risk | Categorical |
| Marital_Status | Marital status | Categorical |
| Food_Insecurity | Food insecurity | Categorical |
| Num_Abortions | Number of abortions | Categorical |
| Num_Liv_Children | Number of living children | Categorical |
| Num_Pregnancies | Number of pregnancies | Categorical |
| Fam_Planning | Family planning information | Categorical |
| Educ_Level | Educational level | Categorical |
| Fam_Income | Family income | Categorical |
| Age | Age of pregnant woman | Numerical |
| Mother_Syphilis_Test_Result | Mother's first syphilis test result | Categorical |

To retrain the model based on the Original Teixeira scenario, we don’t use the Mother_VDRL_Result attribute and apply the random undersampling data balancing technique. This results in a data set containing 48 attributes and 2,004 records with an equal distribution of 1,002 records for each class.

In Table 1, the Mother_VDRL_Result is a categorical attribute with values such as positive, negative, and not informed. We transformed it into binary format (positive and negative). The ‘not informed’ category includes pregnant women who either did not undergo the VDRL exam or rapid test for syphilis, or underwent the exams but their results were not recorded in the system. Consequently, all records associated with ‘not informed’ values in the Mother_VDRL_Result attribute were excluded. This led to a data set comprising 11,201 records, with 344 testing positive for CS and 10,857 testing negative. To tackle the class imbalance, we employed the same data balancing technique as used in the Teixeira model, resulting in a balanced data set for the VDRL model, consisting of 688 records. This data set is utilized for training various machine learning models in the Silva scenario.

The methodology used to train and test all models starts by splitting our data set into training (80%) and testing (20%) sets. The training set was used to train the models, while the test set was reserved exclusively for evaluating the performance of the models in the final stage.

In this paper, we utilize the following tree-based models for analysis: Random Forest, AdaBoost, Gradient Boost, and XGBoost. These models incorporate hyperparameters that play a crucial role in the learning process, but are not learned during the training. To identify the best hyperparameters for training, we employed the grid search technique. This technique involves systematically exploring a predefined search space to evaluate all possible combinations and determine the best learning configuration among the options.

For the grid search, we utilize a k-fold cross-validation, with k=5, and evaluate the performance of each model using the F1-score as the evaluation metric. The search space for each model, specifying the factors and levels, is summarized in Table 2.

**Table 2.** Grid Search factors and levels

| Model | Hyperparameters | Values |
| --- | --- | --- |
| Random Forest | n_estimators | [50, 100, 150, 200] |
|  | criterion | ['entropy', 'gini'] |
|  | max_depth | [None, 1, 3, 5, 7, 9, 11] |
| AdaBoost | n_estimators | [50, 100, 150, 200] |
|  | learning_rate | [0.01, 0.1, 0.5, 1] |
| GradientBoost | n_estimators | [50,100,150,200] |
|  | learning_rate | [0.01, 0.1, 0.5, 1] |
|  | loss | ['deviance', 'exponential'] |
|  | max_depth | [None, 1, 3, 5, 7, 9, 11] |
| XGBoost | learning_rate | [0.01, 0.1, 0.5, 1] |
|  | 'max_depth | [1, 3, 5, 7, 9, 11] |
|  | n_estimators | [50,100,150,200] |

After executing the grid search, we obtain the best hyperparameters for each model. We then proceed to the model evaluation phase. Since the results of our experiments are deterministic, we do not calculate the cut off for significance (p-value) used to conduct statistical analysis, but therefore to quantitatively evaluate the models, we use the following evaluation metrics: accuracy, precision, sensitivity, specificity, and F1-score. Our experiments were performed on Google Colab, using Python with pandas version 1.5.3, numpy version 1.22.4, sklearn version 1.2.2 and xgboost version 1.7.6 libraries.

## Results

The overall baseline characteristics of the pre-processed data set related to the pregnant women assisted by the PMCP are presented in Table 3.

**Table 3.** Baseline characteristics of the data set.

| Variables | Total | Positive | Negative |
| --- | --- | --- | --- |
| Total: n (%) | 11201 | 10857 | 344 |
| Plan_Pregnancy: n (%) |  |  |  |
| Yes | 4606 (41.1) | 4490 (41.4) | 116 (33.7) |
| No | 6517 (58.2) | 6296 (58.0) | 221 (64.2) |
| Missing | 78 (0.7) | 71 (0.7) | 7 (2.0) |
| Has pregnancy risk: n (%) |  |  |  |
| Yes | 1689 (15.1) | 1604 (14.8) | 85 (24.8) |
| No | 9014 (80.5) | 8778 (80.9) | 236 (68.6) |
| Missing | 498 (4.4) | 475 (4.4) | 23 (6.7) |
| Marital status: n (%) |  |  |  |
| Single | 3621 (32.3) | 3495 (32.2) | 126 (36.6) |
| Married | 2624 (23.4) | 2582 (23.8) | 42 (12.2) |
| Widowed | 20 (0.2) | 19 (0.2) | 1 (0.3) |
| Judicial separation | 31 (0.3) | 29 (0.3) | 2 (0.6) |
| Divorced | 78 (0.7) | 77 (0.7) | 1 (0.3) |
| Other | 4827 (43.1) | 4655 (42.9) | 172 (50.0) |
| Food insecurity: n (%) |  |  |  |
| Yes | 4939 (44.1) | 4846 (44.6) | 93 (27.0) |
| No | 1989 (17.8) | 1942 (17.9) | 47 (13.7) |
| Missing | 4273 (38.1) | 4069 (37.5) | 204 (59.3) |
| Number of abortions: n (%) |  |  |  |
| None | 4354 (38.9) | 4233 (39.0) | 121 (35.2) |
| One | 1397 (12.5) | 1335 (12.3) | 62 (18.0) |
| More than one | 335 (3.2) | 344 (3.2) | 11 (3.2) |
| Missing | 5095 (45.5) | 4945 (45.5) | 150 (43.6) |
| Number of living children: n (%) |  |  |  |
| None | 1650 (14.7) | 1604 (14.8) | 46 (13.4) |
| One | 2962 (26.4) | 2883 (26.6) | 79 (23.0) |
| Two | 1359 (12.1) | 1303 (12.0) | 56 (16.3) |
| More than two | 937 (8.4) | 894 (8.2) | 43 (12.5) |
| Missing | 4293 (38.3) | 4173 (38.4) | 120 (34.9) |
| Number of pregnancies: n (%) |  |  |  |
| None | 1070 (9.6) | 1027 (9.5) | 43 (12.5) |
| One | 3558 (31.8) | 3476 (32.0) | 82 (23.8) |
| Two | 2391 (21.3) | 2321 (21.4) | 70 (20.3) |
| More than two | 2536 (22.6) | 2446 (22.5) | 90 (26.2) |
| Missing | 1646 (14.7) | 1587 (14.6) | 59 (17.2) |
| Received information about family planning: n (%) |  |  |  |
| Yes | 5852 (52.2) | 5683 (52.3) | 169 (49.1) |
| No | 3523 (31.5) | 3417 (31.5) | 106 (30.8) |
| Missing | 1826 (16.3) | 1757 (16.2) | 69 (20.1) |
| Level of schooling: n (%) |  |  |  |
| Illiterate | 101 (0.9) | 96 (0.9) | 5 (1.5) |
| Complete elementary school | 306 (2.7) | 294 (2.7) | 12 (3.5) |
| Incomplete elementary school | 1134 (10.1) | 1080 (9.9) | 54 (15.7) |
| Complete middle school | 637 (5.7) | 613 (5.6) | 24 (7.0) |
| Incomplete middle school | 2294 (20.5) | 2217 (20.4) | 77 (22.4) |
| Complete high school | 4003 (35.7) | 3918 (36.1) | 85 (24.7) |
| Incomplete high school | 1942 (17.3) | 1867 (17.2) | 75 (21.8) |
| Complete superior school | 337 (3.0) | 332 (3.1) | 5 (1.5) |
| Incomplete superior school | 225 (2.0) | 223 (2.1) | 2 (0.6) |
| Missing | 222 (2.0) | 217 (2.0) | 5 (1.5) |
| Family income: n (%) |  |  |  |
| Less than or equal to R\$ 500.00 | 4563 (40.7) | 4436 (40.9) | 127 (36.9) |
| Between R501.00 and R 1000.00 | 2293 (20.5) | 2213 (20.4) | 80 (23.3) |
| More than R\$ 1001.00 | 1125 (10.0) | 1090 (10.0) | 35 (10.2) |
| Missing | 3220 (28.7) | 3118 (28.7) | 102 (29.7) |
| Age: mean (SD) | 25.5 (6.6) | 25.5 (6.6) | 25.0 (6.5) |
| Mother's first syphilis test result: n (%) |  |  |  |
| Positive | 428 (3.8) | 208 (1.9) | 220 (64.0) |
| Negative | 10773 (96.2) | 10649 (98.1) | 124 (36.0) |

Table 4 displays the best hyperparameters found by the grid search for each model, along with the evaluation metrics using the test set, for both the Updated Teixeira scenario and the Silva scenario.

**Table 4.**
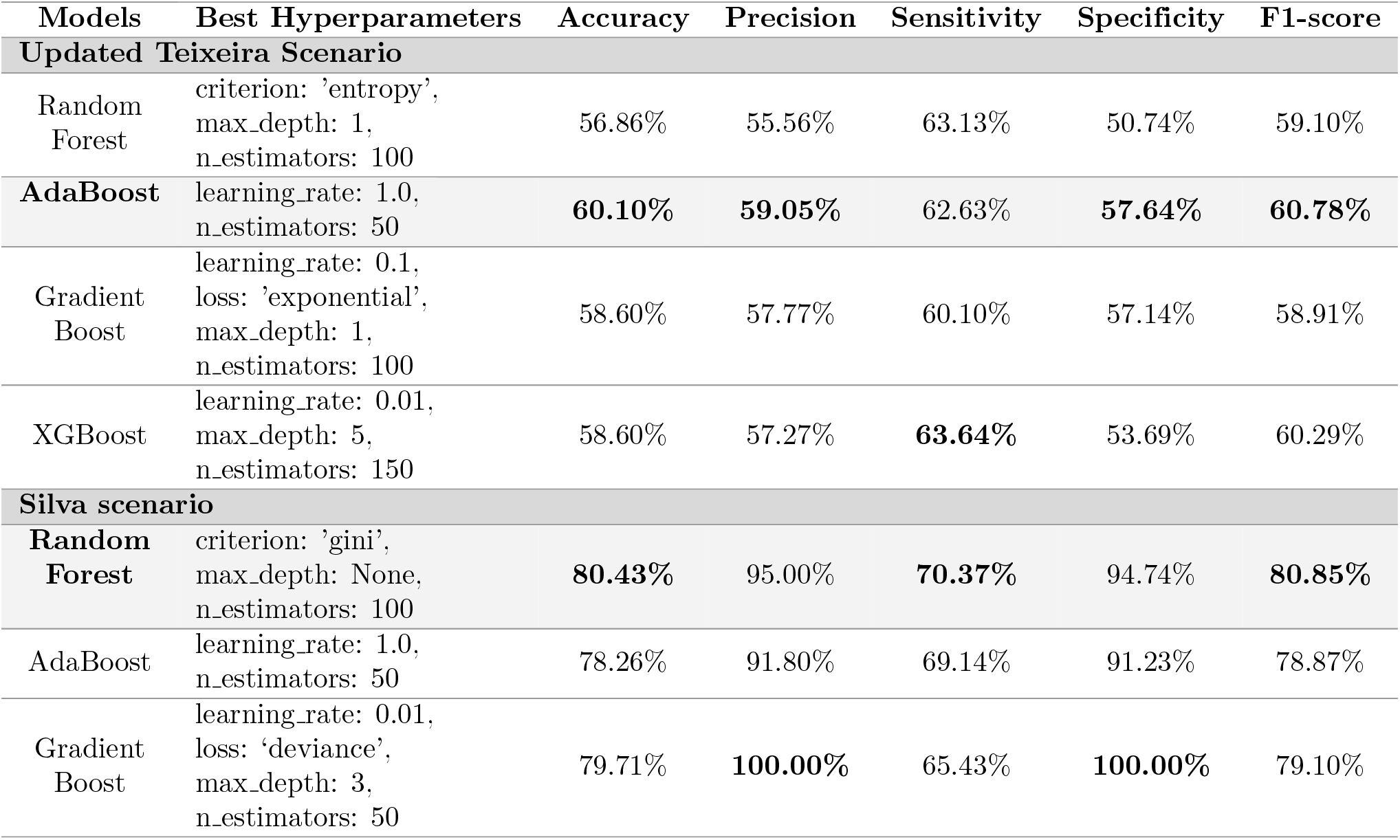
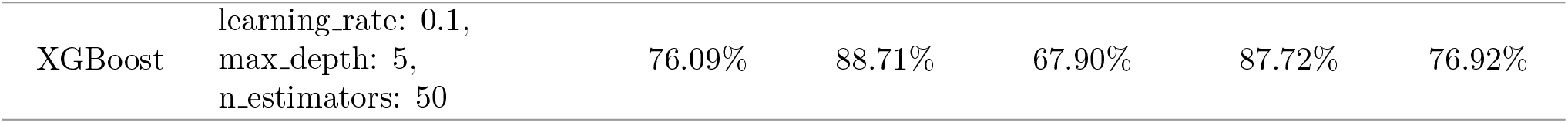
The best hyperparameters from grid search and the respective evaluation metrics for the Teixeira scenario with updated data set and Silva scenario using the test set.

Regarding the models from the Updated Teixeira scenario, their performance is quite similar overall, with AdaBoost generally achieving the best results in terms of accuracy, precision, sensitivity, specificity, and F1-score. The XGBoost had the highest sensitivity at 63.64%, closely followed by Random Forest at 63.13%, indicating their potential for effectively capturing positive instances. Considering the specific implications for predicting CS, the results highlight the importance of models that can strike a balance between high sensitivity and precision.

As for the models from the Silva scenario, the Random Forest achieved the highest accuracy of 80.43%, F1-score of 80.85%, and a sensitivity of 70.37% compared to the other models. Notably, the Gradient Boost exhibited high precision and specificity, but its sensitivity was significantly lower compared to the other models. Additionally, when excluding the Gradient Boost, the Random Forest still demonstrates the best performance in terms of precision (95%), and specificity (94.74%). These results highlight the robustness and effectiveness of the Random Forest within the Silva scenario for predicting CS.

In order to compare the results obtained, we designate Adaboost as the best model for the Updated Teixeira scenario, calling it the **Updated Teixeira model**. For the Silva scenario, the Random Forest model was identified as the best choice, referred to as the **Silva model**. Figure 3 presents a radar graph where we show the Updated Teixeira model, the Silva model and the Original Teixeira model based on the five evaluation metrics.

**Fig 3.**
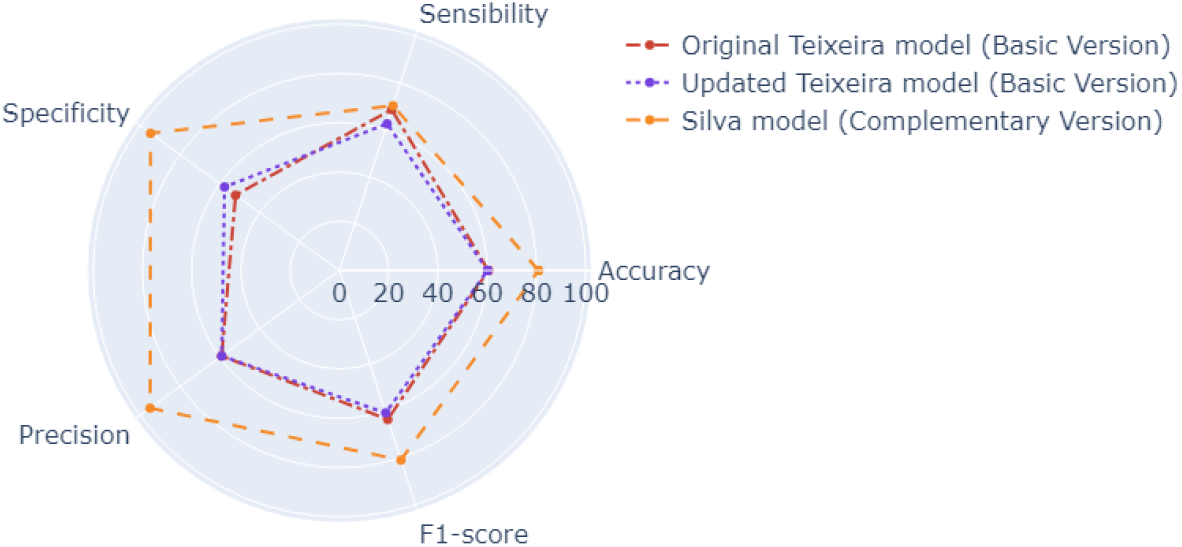
Radar graph comparing the best models of each scenario.

These findings suggest that the inclusion of additional information about the Mother_VDRL_Result led to an enhancement in the model’s ability to accurately identify negative cases (improved specificity) while slightly compromising its ability to correctly identify positive cases (decreased sensitivity). Overall, the Silva model demonstrated superior performance compared to the Original Teixeira models across all metrics. There was an improvement in the specificity metric, increasing from 52.12% to 94.74%. However, the model still encountered challenges in terms of sensitivity, with an increase from 68.67% to 70.37%.

## Discussion

The main objective of this study was to examine whether the inclusion of additional information acquired during the gestational follow-up process leads to an improvement in the predictive performance of models for CS. In this respect, our results confirm that the proposed approach, the Silva scenario, consistently outperforms the benchmark models across multiple machine learning algorithms (Random Forest, AdaBoost, Gradient Boost, and XGBoost). The Silva models outperform the benchmarks across all metrics including accuracy, precision, sensitivity, specificity, and F1-score, indicating its effectiveness in predicting CS, and as such providing greater predictive power and valuable insights for early intervention and appropriate medical care. Furthermore, we confirm our hypothesis regarding the importance of the inclusion of the Mother_VDRL_Result in the prediction of CS using machine learning.

From a practical perspective, both options are not mutually exclusive. Both models can be used as decision support tools for healthcare professionals depending on the local circumstances. For example, the Original Teixeira model can be employed for all pregnant women, offering crucial signals to healthcare professionals for required and improved treatment without the need for additional laboratory tests and associated delays. However, when and if a pregnant woman undergoes the first VDRL exam or a rapid test for syphilis, it might be advisable to utilize the Silva model, which exhibits stronger evaluation metrics. This twin track approach ensures the utilization of both models to support healthcare decisions and enhances the precision of predictions for pregnant women at different stages of their pregnancy.

There are a number of strengths and limitations to this study. The data is longitudinal, current, and relatively large for this type of study and compares favorably with prior research. As is evident from the results, despite the number of cases of CS in the sample being low, the proposed approach resulted in acceptable performance.

Notwithstanding this, larger and more diverse cohorts from across Brazil and other countries would likely result in even better performance particularly with respect to sensitivity. While over time, increased digitisation combined with improved record keeping will result in larger data sets in Brazil, in the near future, greater use of international data is anticipated.

As discussed earlier, in Brazil, the volume of empty records in the SIS-MC is a significant issue and a substantial percentage of people with syphilis are asymptomatic. Treating syphilis as early as possible can significantly reduce complications for mothers and babies. The effective operationalisation of machine learning-based decision support tools, such as the Original Teixeira model and Silva model, requires the required data being collected as completely, accurately and early as possible. Relatedly, new data attributes may also result in greater prediction accuracy particularly in the case of machine learning models that rely on attributes chosen by health experts such as the BODS-Expert approach used in Teixeira et al. [1] and this study. For example, this may include the number of prenatal care appointments, clinical data on partners, settlement type (urban, rural, or sparse settings) as well as other sociodemographic indicators. This infers greater collaboration is required between healthcare professionals and machine learning researchers and designers.

This study has achieved high predictive values for CS. Firstly, it is unclear whether similar approaches can be followed for manifestations of syphilis where data on the focal person may differ from that collected for pregnant women. Secondly, prognosis is not considered which may prove to be a fruitful and impactful avenue of research.

In conclusion, machine learning models with sufficient clinical and administrative (incl. sociodemographic) data can present promising results for predicting the risk of CS. With greater, more diverse, and more complete data, model performance and in particular model sensitivity can be improved even further. Providing individualized probabilities of CS to healthcare practitioners as a decision support tool can have a dramatic impact on reducing complications for mothers and babies. Unfortunately, the highest incidences of CS are often those countries where there are substantial issues in relation to digital health literacy and the routine collection and availability of relevant data. Challenges which must be overcome if the promised positive impacts of machine learning in health is to be fulfilled.

## Conclusions

The significant surge in reported cases of CS underscores a mounting public health challenge in Brazil [12]. While tests and treatments for syphilis are accessible through primary health care, early detection of these risks during pregnancy is crucial for effective treatment and monitoring, thereby averting adverse outcomes. In this work, investigated and compared Machine Learning models for categorizing CS, utilizing clinical and sociodemographic data factors, and assessing the model’s efficacy in incorporating additional attributes during pregnancy.

We evaluated four machine learning techniques: Random Forest, AdaBoost, Gradient Boost, and XGBoost. We used the BODS Experiment proposed in Teixeira et al. [1], which in the work in question obtained the best results. We selected the same 11 attributes selected by this work, and added information from the mother’s first syphilis test (either the VDRL exam or rapid test).

Our results demonstrated the gain in the performance of the models by adding new information that is collected during the gestational period, improving all the analyzes evaluated. It is worth noting that a large amount of missing data may have impacted model learning, reducing the need for PMCP to improve the quality of data acquisition.

Finally, it is important to understand that this work does not aim to replace the models presented in Teixeira et al. [1], but that together, they can more positively assist healthcare professionals in decision-making. As future work, we intend to apply different techniques to deal with missing data, and propose a tool that integrates the models presented in this work.

## Data Availability

All data are available from the Mendeley Data database (accession number(s) doi:10.17632/3zkcvybvkz.2).

https://www.doi.org/10.17632/3zkcvybvkz.2

## Acknowledgments

This work was funded by Conselho Nacional de Desenvolvimento Científico e Tecnológico (CNPq), Coordenação de Aperfeiçoamento de Pessoal de Nível Superior (CAPES), Fundação de Amparo à Ciência e Tecnologia do Estado de Pernambuco (FACEPE), and Universidade de Pernambuco (UPE). Patricia Takako Endo is funded by CNPq via PQ 2 grant.

